# Effect of Tocilizumab on “ventilator free days” composite outcome in SARS-CoV-2 patients. A retrospective competing risk analysis

**DOI:** 10.1101/2021.04.01.21254794

**Authors:** Ahmed F. Mady, Basheer Abdulrahman, Omar E. Ramadan, Shahzad A. Mumtaz, Mohammed A. Al-Odat, Ahmed Kuhail, Rehab Altoraifi, Rayan Alshae, Abdulrahman M. Alharthy, Dimitrios Karakitsos, Waleed Th. Aletreby

**Author notes:** Corresponding author: Waleed Th. Aletreby.

## Abstract

**Background:** SARS-CoV-2 infection demonstrates a wide range of severity, the more severe cases demonstrate a cytokine storm with elevated serum interleukin-6, hence IL-6 receptor antibody Tocilizumab was tried for the management of severe cases.

**Objectives:** The effect of Tocilizumab treatment on the composite outcome of ventilator free days, among critically ill SARS-CoV-2 patients.

**Method:** Retrospective observational propensity score matching study, comparing mechanically ventilated patients upon ICU admission who received Tocilizumab to a control group. Utilizing competing risk analysis method, and reporting sub-distributional hazard ratio of a composite outcome of ventilator free days at day 28.

**Results:** 29 patients in the intervention group were compared to 29 patients in the control group. Matched groups were similar at base line. The primary outcome of ventilator free days was higher in the intervention group (SHR 2.7, 95% CI: 1.2 – 6.3; p = 0.02), crude ICU mortality rate was not different between Tocilizumab and control groups (37.9% versus 62% respectively, p = 0.1), actual ventilator free days were significantly longer in Tocilizumab group (mean difference 4.7 days, 95% CI 1.1 – 8.3; p = 0.02). Sensitivity analysis by Cox regression showed a significantly lower hazard ratio of death in Tocilizumab group (HR 0.49, 95% CI: 0.25 – 0.97; p = 0.04). While there was no difference in grown positive cultures among groups (55.2% in Tocilizumab group versus 34.5% in the control, 95% CI of difference: −7.11% to 54.4%; p = 0.1).

**Conclusion:** Tocilizumab may improve the composite outcome of ventilator free days at day 28 among mechanically ventilated SARS-CoV-2 patients, it is associated with significantly longer actual ventilator free days, and insignificantly lower mortality and superinfection.

## Introduction

An international pandemic was declared by the World Health Organization (WHO) on March 11^th^, 2020 as result of a rapidly spreading viral infection causing pneumonia and respiratory symptoms ^(1)^, the responsible virus was identified to be a novel positive-sense-single strand RNA virus, of the family coronaviridae, capable of infecting a range of hosts including humans, that soon came to be recognized as SARS-CoV-2 ^(2)^. Despite all measures of containment, the pandemic spread all over the world, so that by October 15^th^, 2020 more than 38 million cases have been confirmed, and more than 1 million fatalities worldwide ^(1)^. It is well known, however; that not all positive cases demonstrated similar signs and symptoms, nor severity. In fact, SARS-CoV-2 infection demonstrates a fairly wide spectrum of symptoms, ranging from asymptomatic cases, to mild respiratory complains, to severe pneumonia, up to severe acute respiratory distress syndrome (ARDS) ^(3)^, up to 10% of cases are on the severe end of the spectrum, suffering in addition to ARDS multi-organ failure, and requiring mechanical ventilation and/or admission to intensive care units (ICU) ^(4)^.

Critically ill patients with SARS-CoV-2 infection showed elevated levels of pro-inflammatory cytokines, particularly interleukin 2 and 6 (IL2, IL6) among others ^(5)^, serum levels of such cytokines -particularly IL6 -were higher compared to patients with milder presentation ^(6)^, and have been associated with increased mortality and poor outcomes ^(6,7)^. Such findings, coupled with post-mortem evidence of severe alveolar edema, patches of inflammatory infiltrates, and proteinaceous exudates ^(8)^, suggest that a cytokine storm secondary to dysregulated immune response of the host may be associated with SARS-CoV-2 infection ^(4, 9)^.

Tocilizumab (TCZ) is a humanized anti-interleukin 6 receptor monoclonal antibody ^(5)^, that targets both forms of receptors namely soluble and membrane bound receptors ^(10)^, accordingly it has been postulated that TCZ treatment may be able to attenuate the so called “cytokine storm” associated with SARS-CoV-2 infection, and prevent the progress of the infection into ARDS ^(4, 5)^, and it was licensed by the United States Food and Drug Administration (FDA) for the management of cytokine release syndrome ^(11)^. Hence, this study was performed to assess the impact of treatment with TCZ on critically ill patients with SARS-CoV-2, utilizing a composite outcome measure that is commonly used in critical care studies, ventilator free days (VFD) at 28 days is used to quantify the efficacy of therapies and interventions on morbidity in the presence of the competing event of death ^(12, 13)^. Our hypothesis was that treatment with TCZ would reduce the composite outcome of VFD.

### Objectives

The primary objective was 28 days VFD as a composite outcome, while secondary outcomes included components of VFD composite outcome, namely, intensive care unit (ICU) mortality and actual ventilator free days (aVFD), in addition to positive grown cultures as an adverse event.

## Method

### Study Design

This was a retrospective observational study that exploited analytical statistical methods to compare the outcomes of mechanically ventilated patients who received TCZ to those who didn’t during SARS-CoV-2 pandemic, as a composite outcome of both mortality and duration of mechanical ventilation.

### Setting and timeframe

This study was conducted in the adult ICU of King Saud Medical City (KSMC), the largest Ministry of Health hospital in the central region of Saudi Arabia. The ICU harbors originally 127 beds, and was expanded during SARS-CoV-2 pandemic to include 300 beds, half of which were isolation single rooms, and the rest were open cohorting areas. It is a closed ICU operated 24/7 by dedicated intensivists, with a nursing ratio of 1:1. During the SARS-CoV-2 pandemic KSMC was a COVID referral center, and we generally followed management protocols recommended by the Ministry of Health, which are an adaptation of international guidelines. This study included patients admitted to the ICU in the period between March 1^st^, 2020 and June 30^th^, 2020. The analysis included a subset of patients from a previously published article ^(3)^.

Inclusion criteria: we included patients if they were admitted to the ICU during the study period, adults (age ≥ 18), mechanically ventilated upon ICU admission, confirmed COVID-19 positive (by RT-PCR nasopharyngeal swab) and received two doses of TCZ during the course of their treatment. TCZ is given in our ICU in the dose of 4-8 mg/kg, as an intravenous infusion reconstituted in 100 ml of 0.9% sodium chloride solution over 60 minutes.

We excluded patients younger than 18 years old, pregnant females, and known pulmonary tuberculosis and human immunodeficiency virus (HIV) positive cases. Data of all other patients with the same criteria (apart from receiving TCZ) could be used to identify a control group (further details follow).

The original study was approved by the local institutional review board (IRB) with waiver of consent in view of its retrospective design, both studies observe the general principals outlined by the declaration of Helsinki.

### Data management

We retrieved demographic data of all patients who fulfilled the inclusion criteria (age, gender, comorbidities, smoking status, severity score, and body mass index), all included patients must have been mechanically ventilated upon ICU admission, and we recorded the date of extubation (if at all) within 28 days, and the ICU outcome as a binary variable of death or survival. Furthermore, we recorded other modalities of treatment including anti-viral, and steroids. Finally, we noted reports of positive cultures grown during ICU stay.

### Outcomes

The primary outcome was a composite outcome of VFD, it is a common outcome frequently utilized in clinical trials in ICU to quantitatively explore the effect of an intervention or treatment on morbidity in the presence of the competing risk of death ^(12, 13)^, details of VFD are given elsewhere ^(14)^, briefly; if the patient within 28 days dies or remains intubated, the outcome is considered a failure, and the patient is awarded zero aVFD. The outcome is considered a success only if the patient was extubated before 28 days, and was still alive at day 28, in such case the aVFD are the days between extubation and day 28. The outcome doesn’t award aVFD if the patient was re-intubated, or died within 28 days after being extubated. Secondary outcomes included ICU mortality, and aVFD, in addition to grown cultures (of any source or organism) as an adverse event that may arise due to the use of TCZ.

### Statistical methods

Patients who fulfilled the inclusion criteria constituted the intervention group, and we used the data of all other patients with the same criteria apart from receiving TCZ to identify a control group, using propensity score matching, we intuitively chose matching criteria of: age, gender, severity score, comorbidities, smoking status, body mass index (BMI), and the receipt of steroids and antiviral medications. Matching was 1:1 nearest neighbor method with a caliber width of 0.2 without replacement. The reason we didn’t follow the classical method of propensity score matching where logistic regression is performed using receiving TCZ as the dependent to identify variables to match upon ^(15)^ is that we expected a small number of patients receiving TCZ with numerous matching criteria so that if all were included in a logistic regression model would have violated the rule of thumb of 10 events / variable and that would have resulted in over fitting ^(16)^.

Once the matched control group was identified the primary outcome was assessed in a competing risk regression analysis, utilizing the patients’ status as alive and extubated as the event of interest, whereas dead or still intubated as the competing risk ^(14)^, the primary outcome was reported as sub-distribution hazard ratio (SHR) according to Fine and Gray method ^(17)^, in this method the risk of interest and the competing risk are mutually exclusive, that is to say if one event occurred, the other can’t.

We summarized data of the intervention and control groups by median and interquartile range (IQR) for continuous variables, and compared them by student t-test or Wilcoxon rank sum test as appropriate. We summarized categorical variables as frequency and percentage, and compared them by chi^2^ or Fisher’s exact tests as appropriate. Comparisons were presented with corresponding 95% confidence interval (CI) and p values.

Furthermore, we planned apriori to compare 28-day survival among both groups (regardless of the mechanical ventilation status) in a Cox Proportional Hazard regression model as a sensitivity test for the primary outcome, the result of which we presented as p value of log rank test, along with the corresponding Kaplan Meier curve.

All statistical tests were two sided, considered statistically significant with p value < 0.05, without correction for multiple testing. Commercially available software STATA® was used in the analysis (StataCorp. 2017. *Stata Statistical Software: Release 15*. College Station, TX: StataCorp LLC.).

## Results

During the study period seven hundred forty two SARS-CoV-2 positive patients were admitted to the ICU, out of those four hundred sixty seven patients were intubated and mechanically ventilated upon ICU admission, we further excluded another fifty two patients (46 younger than 18 years old, 2 pregnant females, 3 known pulmonary tuberculosis, and one known HIV case), we screened the remaining four hundred fifteen patients and identified 29 patients who received TCZ according to our ICU protocol, those constituted the intervention group. Out of the three hundred eighty six patients who didn’t receive TCZ we were able to match 29 patients to constitute the control group through the previously described propensity score matching method (Figure 1). Comparison of the demographic and clinical management characteristics of the intervention group to the unmatched group of mechanically ventilated patients showed imbalances in age and sequential organ failure assessment (SOFA) scores upon ICU admission, both variables being significantly lower in the unmatched group, those imbalances were corrected after propensity score matching, and the intervention and control groups had no statistically significant differences (Table 1).

**Table 1:** Demographic and clinical characteristics of study groups before and after matching:

| Variable | Unmatched Groups |  |  | Matched Groups |  |  |
| --- | --- | --- | --- | --- | --- | --- |
|  | TCZ (n=29) | No TCZ (n=386) | P value | TCZ (n = 29) | No-TCZ (n = 29) | P value |
| Age: Median (IQR) | 59 (52.5 – 60.25) | 51 (44 – 57) | 0.0001 | 59 (52.5 – 60.25) | 57 (51.75 – 63.25) | 0.9 |
| Gender: Male (n, %) | 21 (72.4%) | 262 (67.9%) | 0.6 | 21 (72.4%) | 20 (69%) | 0.8 |
| SOFA score Median (IQR) | 4 (4 – 5) | 3 (4 – 5) | 0.0001 | 4 (4 – 5) | 4 (4 – 4.25) | 0.2 |
| BMI Median (IQR) | 27 (23.3 – 31.2) | 25.8 (22.6 – 29.4) | 0.2 | 27 (23.3 – 31.2) | 25.3 (21.85 – 28.63) | 0.1 |
| Smoking (n, %) | 10 (34.5%) | 131 (33.9%) | 0.9 | 10 (34.5%) | 9 (31%) | 0.8 |
| Anti-viral (n, %) | 11 (37.9%) | 154 (39.9%) | 0.8 | 11 (37.9%) | 12 (41.4%) | 0.8 |
| Steroids (n, %) | 17 (58.6%) | 245 (63.5%) | 0.6 | 17 (58.6%) | 16 (55.2%) | 0.8 |

**Figure 1:**
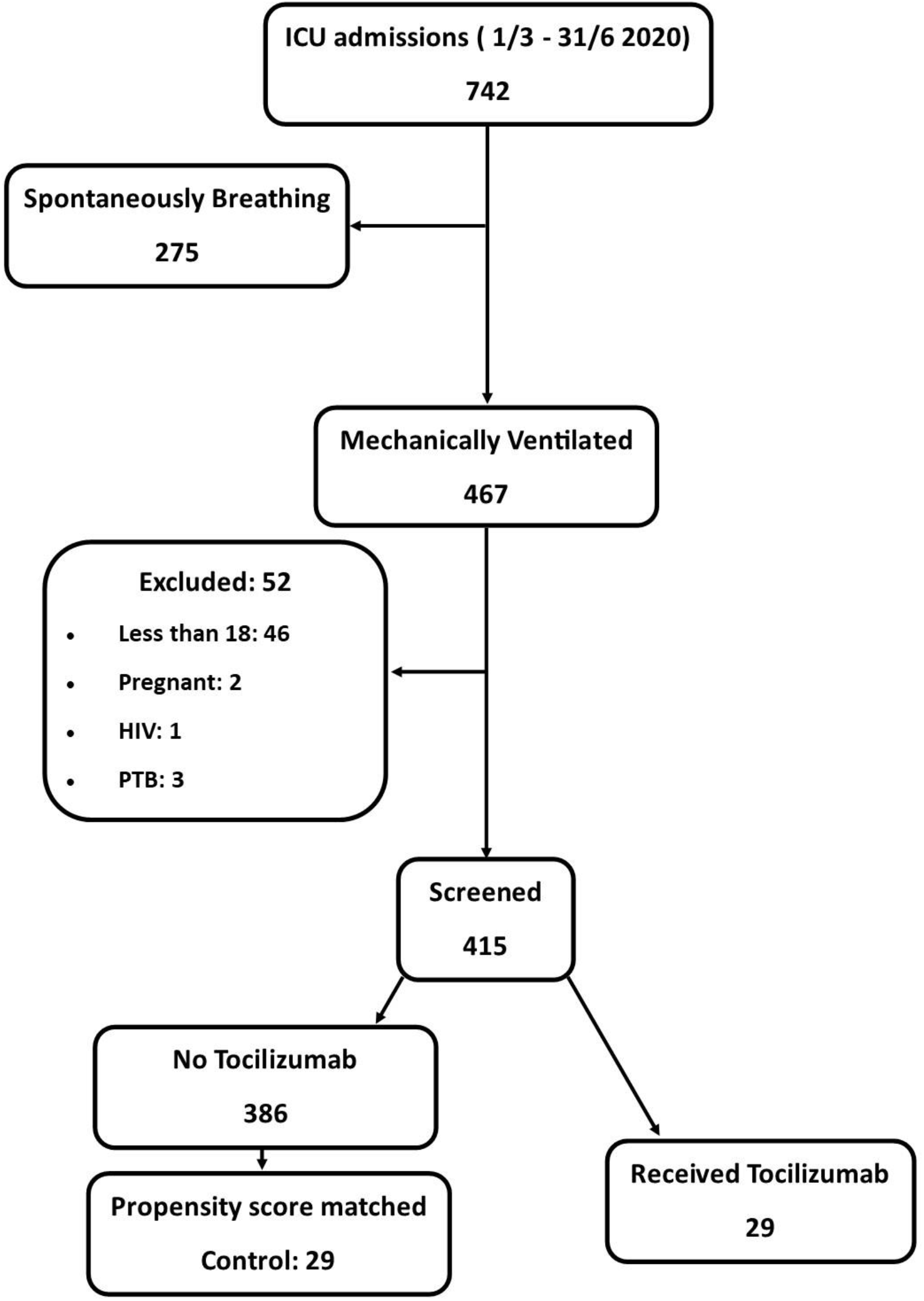
Patients’ and study groups’ flow diagram:

The primary outcome of our study was the SHR of being alive and extubated at 28 days as the outcome of interest, in the presence of the competing risk of death or still intubated at 28 days, our analysis revealed that receiving TCZ results in a statistically significant SHR of 2.7 (95% CI: 1.2 – 6.3; p = 0.02), that is to say increasing the proportional “hazard” of being alive and extubated at day 28 by 170% compared to patients who didn’t receive TCZ. Our sensitivity analysis in the form of Cox regression supports our findings, the HR of the Cox regression model (for the hazard of death) was 0.49 (95% CI: 0.25 – 0.97; log rank p = 0.04). In agreement with the primary outcome, the result of Cox regression indicates that receiving TCZ reduces the relative hazard of death by 51% (Table 2). The Kaplan Meier survival curve of both groups is depicted in figure 2.

**Table 2:** Results of competing risk analysis, and Cox proportional regression:

|  | Sample statistic (95% CI) | P value |
| --- | --- | --- |
| Competing Risk Regression | SHR: 2.7 (1.2 – 6.3) | 0.02 |
| Cox Proportional Hazard | HR: 0.49 (0.25 – 0.97) | 0.04 |

**Figure 2:**
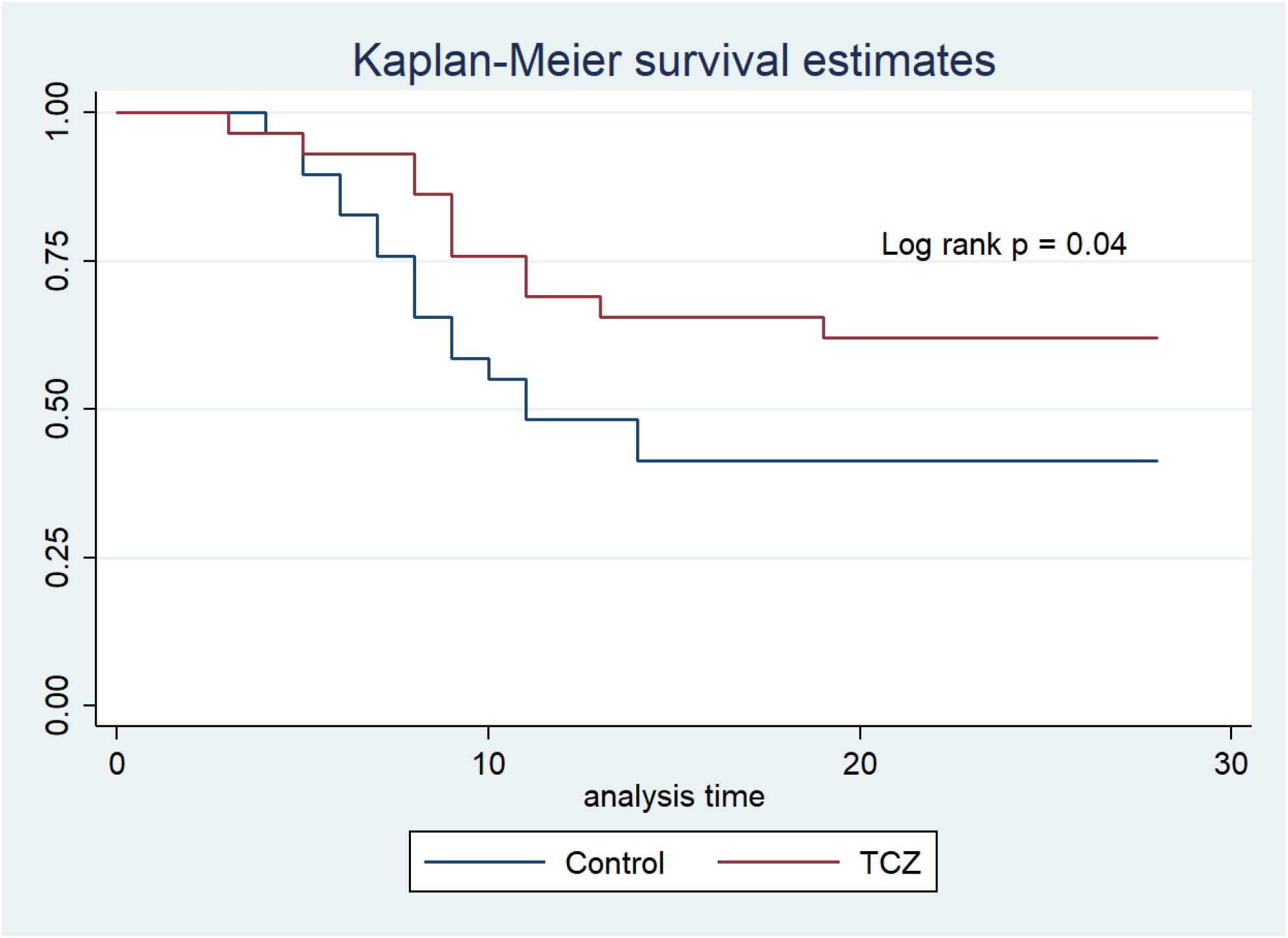
Kaplan Meier survival curve:

The secondary outcomes show that 11 (37.9%) patients in the TCZ group died within 28 days, while 18 patients (62%) of the control group died within the same period, although numerically lower the difference in mortality rate between groups was not statistically significant (95% CI of difference: −3.9% to 48.5%; p = 0.1), all occurrences of death took place in the ICU and none of the patients was spontaneously breathing at the time of death. The aVFD, however; was statistically higher in the TCZ group, median (IQR) of aVFD in TCZ group was 10 (0 – 13) compared to 0 (0 – 2.25) in the control group (mean difference 4.7, 95% CI of difference: 1.1 – 8.3; p = 0.02). In the TCZ group 16 patients (55.2%) have grown positive cultures during the study period, whereas, only 10 patients (34.5%) in the control group did. The higher rate in TCZ group was not statistically significant (95% CI of difference: −7.11% to 45.4%; p = 0.1). Table 3 shows the results of secondary outcomes.

**Table 3:** Results of secondary outcomes:

|  | TCZ (n=29) | Control (n=29) | 95% CI of difference | P value |
| --- | --- | --- | --- | --- |
| ICU mortality (n, %) | 11 (37.9%) | 18 (62%) | -3.9 to 48.5 | 0.1 |
| Actual VFD: median (IQR) | 10 (0 – 13) | 0 (0 – 2.25) | 1.1 – 8.3 | 0.02 |
| Positive cultures (n, %) | 16 (55.2%) | 10 (34.5%) | -7.11 to 45.4 | 0.1 |

## Discussion

In this study 29 mechanically ventilated SARS-CoV-2 positive patients who received TCZ were compared to a propensity score matched control group of 29 patients with similar characteristics, the primary outcome was the composite outcome VFD at day 28, it encompasses both duration of mechanical ventilation and mortality. In our competing risk analysis, receiving TCZ harbored a statistically significant higher SHR (2.7, 95% CI: 1.2 – 6.3; p = 0.02) of being alive and extubated at 28 days, compared to those who didn’t receive TCZ. This type of outcome is common in trials of ARDS in critically ill patients ^(18-20)^ due to several advantages, for example it penalizes mortality making it a plausible trial endpoint, while including the continuous variable of ventilator days adds to statistical power, in addition to being realistic since a single intervention in ARDS is unlikely to impact mortality, however; it may shorten the duration of ventilation if it improves the lung condition, because – as is the case in SARS-CoV-2 infection-ARDS is heterogeneous and mortality is usually multi-factorial ^(14)^.

To our best knowledge no other study has performed similar competing risk analysis on critically ill SARS-CoV-2 patients receiving TCZ, however; several studies have demonstrated similar patterns to our findings, particularly with regards to the components of the primary outcome. In our study there was an insignificantly lower mortality rate among patients receiving TCZ, this was also shown by Klopfenstien et al ^(21)^ in a retrospective case control study with a similar sample size, however; studies with larger sample sizes were able to demonstrate a statistically significant lower mortality rates for patients treated with TCZ ^(22, 23)^, which may imply a true clinical effect on mortality that we were not able to demonstrate statistically due to lack of power. The second component of the composite outcome (aVFD) in our study was significantly higher in the TCZ group, confirming our intuition of improving lung condition thus hastening extubation, which was also demonstrated by others ^(24)^.

More recent randomized clinical trials (RCT) explored the outcome of ventilator free days as well, although in different statistical models. In REMAP-CAP ^(25)^ report of the immune module using Bayesian statistics, respiratory support-free days in TCZ group had a significantly higher OR compared to control, with > 99.9% of being superior to control. On the contrary, mean and median ventilator days were not different between TCZ and control groups in two other RCTs ^(26, 27)^.

The sensitivity analysis in our study seems to be in support of the result of the primary outcome, and in concordance with many published articles with different designs, showing a statistically significant HR of survival in Cox regression, and significant log rank p values for TCZ treated patients ^(5, 22, 23, 28)^.

Our adverse event measure was growth of positive cultures of any source and organism, which we expected in view of the immunomodulatory effect of TCZ, indeed there was a higher rate in TCZ group, however; it didn’t reach the level of statistical significance. Yet again, in studies with larger sample size this higher rate of superinfection in TCZ group was shown to be significant ^(23, 28)^ in observational studies, RCTs – however-didn’t show a significant difference between groups in rate of super-added infection ^(26,27)^.

Our study suffers several limitations, first there is the limitation inherent within the retrospective design and lack of prospective randomization, although propensity score matching partially compensates this defect. Second, the small sample size in our study definitely renders it underpowered, and consequently significant findings to be idea generating and should be interpreted cautiously. Third, we didn’t follow the classical method of propensity score matching, however; this was for justifiable reasons, and the resultant matched groups were similar. Last, several details were overlooked in our study, such as the duration of symptoms before hospitalization, mechanical ventilation, and TCZ treatment, since the main focus of the study was the duration of mechanical ventilation itself and ICU outcome, and those details were published elsewhere ^(3)^.

## Conclusion

Mechanically ventilated SARS-CoV-2 patients treated with TCZ may have a better composite outcome of ventilator free days, TCZ may be associated with significantly longer actual ventilator free days, but insignificantly lower mortality and superinfection at day 28. Treatment with TCZ may also be associated with better 28 day survival. These findings need to be confirmed by larger prospective randomized trials.

## Data Availability

Data may be requested from the corresponding author by e-mail.

## Acknowledgment

The authors would like to extend their gratitude to the database team of the ICU at KSMC for their efforts in data collection and abstracting.

